# An AI Competency Framework for Emergency Medicine: A Multiphase Consensus Process

**DOI:** 10.64898/2026.09.15.26363099

**Authors:** Carl Preiksaitis, Michael Makutonin, Dania Abu-Jubara, Wan-Tsu Wendy Chang, Robert Cooney, Deborah Diercks, Neehar Kundurti, Brian Kwan, Rachel Liu, Michael Lozano, Cassandra Mackey, Mona Moukaddem, Elspeth Pearce, Jessica Pelletier, Marta Rowh, R. Andrew Taylor, Christian Rose

**Author notes:** Carl Preiksaitis and Michael Makutonin contributed equally to this work and share first authorship. Corresponding author: Carl Preiksaitis, MD, MEd;.

## Abstract

Artificial intelligence (AI) is increasingly embedded in U.S. emergency department workflows, but no medical specialty has defined what competent use of these tools requires of its physicians. Existing accreditation requirements are silent on AI, and parallel national efforts span the learning continuum but are intentionally specialty-agnostic. As one output of a working group of the 2026 Society for Academic Emergency Medicine (SAEM) Artificial Intelligence Consensus Conference, we developed an emergency medicine-specific framework for AI competency. We employed a multiphase consensus design: a Nominal Group Technique session generated themes from a curated set of clinical AI scenarios, and a two-round modified Delphi process (panel n = 13 across 12 academic medical centers), separated by a consolidation videoconference, evaluated and extended the framework. A priori consensus required ≥70% of panelists rating ≥4 on a 5-point Likert scale (≥75% for themes) with interquartile range ≤1; we followed established Delphi reporting guidance. The framework comprises 5 themes (Communicating about AI, Understanding appropriate use cases, Interacting with AI, AI risk management, Cognitive impacts of AI), 5 derived competencies (one-to-one theme-to-competency mapping endorsed by 12 of 13 panelists), 19 subthemes, and 10 retained clinical scenarios. All themes, derived competencies, and rated subthemes met consensus thresholds; 9 of 10 scenarios reached consensus, with 1 retained as a future-state operational model. Two cross-cutting conceptual frames emerged: pre-emptive versus post-hoc AI integration, and tiered competencies as an articulated need rather than a pre-specified answer. This is, to our knowledge, the first specialty-specific AI competency framework for a United States medical specialty with quantitative content validity evidence from expert consensus. It provides a structural target for emergency medicine curriculum development, assessment design, and faculty development, with downstream priorities including stage-specific competency assignment, assessment instrument development, and periodic re-evaluation as AI deployment evolves.

## Introduction

Artificial intelligence (AI) is now embedded in the workflows of U.S. emergency departments (EDs). AI systems triage patients,^1^ flag life-threatening pathologies on imaging,^2^ predict sepsis and clinical deterioration in real time,^3^ generate discharge instructions,^4^ and, through ambient scribes, document a growing share of clinical encounters.^5^ Emergency medicine (EM)-related AI publications have approximately doubled year-over-year since 2018.^6^ Three-stage developmental frameworks^7^ and practical use-case catalogs^8^ document where the technology has arrived, but neither tells emergency physicians what they need to know to use it safely. A recent precision-emergency-medicine framework called explicitly for “health data literacy” as an educational competency that “all emergency physicians will need training to demonstrate”, without specifying what such a competency contains.^9^

Current EM literature recognizes the potential of AI in teaching and assessment, including adaptive simulation, intelligent tutoring systems, and natural language processing approaches that quantify resident clinical exposure,^10–13^ and highlights the need for more emergency physicians trained in Clinical Informatics to support safe AI integration.^14^ Yet emergency medicine encounters AI under conditions that generalist competencies do not anticipate—undifferentiated, high-acuity patients; compressed decision timelines and diagnostic uncertainty; and elevated medico-legal exposure—often at the point of care rather than in curated workflows. These conditions make an EM-specific educational response necessary, and no publication has yet defined the relevant competencies or outlined how to teach and assess how EM physicians interact with and use AI tools in the clinical environment.

The foundational documents that might be expected to define those competencies are silent. The Accreditation Council for Graduate Medical Education (ACGME) Program Requirements contain no mention of AI or machine learning in any specialty,^15^ and despite rapid growth of AI content in undergraduate curricula, a recent scoping review found neither a standardized approach nor consensus on AI competencies.^16^ The Association of American Medical Colleges (AAMC) is developing competencies that span the learning continuum for release in fall 2026, but these are intentionally specialty-agnostic and do not address discipline-specific needs.^17^ A six-domain AI competency framework for health care professionals offers a profession-wide baseline,^18^ but no specialty has translated such baselines into its own clinical context. EM therefore sits in a space where the regulatory documents do not exist, the national effort is intentionally generic, and no specialty has yet published a consensus competency framework for its workforce. Without one, AI training in EM will continue to develop locally and idiosyncratically, without a shared standard against which to align curricula, assessment, or faculty development.

We met at the 2026 Society for Academic EM (SAEM) AI Consensus Conference, as part of a working group focused on AI education, training, and competency development. Alongside generating prioritized research questions for the conference’s broader research agenda, our group conducted a parallel process to develop an EM-specific AI competency framework. In this article, drawing on a nominal group technique and a two-round modified Delphi, we present the framework, situate it relative to existing competency work in adjacent specialties, and discuss its implications for EM training, assessment, and faculty development.

## METHODS

### Study design and oversight

We used a multiphase consensus design comprising a nominal group technique (NGT) for inductive theme generation, followed by a modified Delphi process incorporating two anonymous online rounds separated by a virtual consolidation meeting (Figure 1).^19,20^ A subsequent post-Round-2 refinement phase, in which the working group co-leads applied a small set of editorial revisions based on panel qualitative feedback, completed the framework. The Delphi component was designed and reported in accordance with the Guidance on Conducting and REporting DElphi Studies (CREDES).^21^ This sequential structure has precedent in digital health competency development^22^ and prior AI competency work.^18^ The study was reviewed by the Institutional Review Board at Stanford (#80201) and determined to be exempt from formal oversight. Additional methods detail is available as supplemental material accompanying the online article (Methods S1).

**Figure 1.**
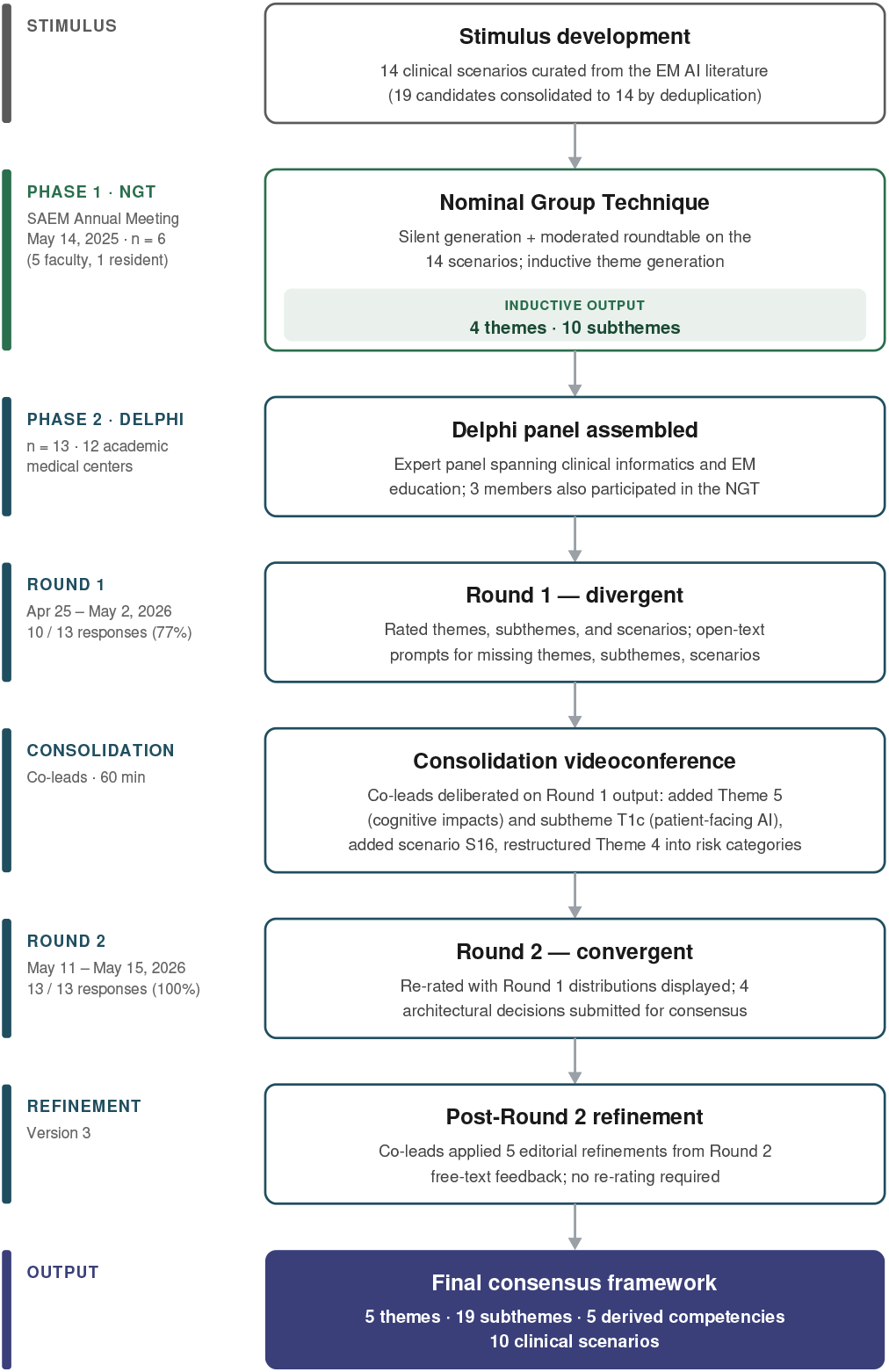
Consensus development process. Themes were generated inductively using a nominal group technique and refined through a two-round modified Delphi with an expert panel (n = 13). The final consensus framework comprises 5 themes, 19 subthemes, 5 derived competencies, and 10 clinical scenarios. Reporting follows CREDES guidance. NGT, nominal group technique; SAEM, Society for Academic Emergency Medicine; CREDES, Conducting and REporting of DElphi Studies.

### Stimulus development

To anchor the consensus process in contemporary EM practice, the working group leadership team developed a set of clinical AI scenarios drawn from the existing literature on AI in emergency care, selected to span the ED patient journey from triage through disposition and to reflect AI applications already implemented or near implementation in United States EDs. Nineteen candidate scenarios entered the NGT and were consolidated to 14 through deduplication of overlapping cases; a fifteenth, covering ambient AI scribe documentation, was added prior to the Delphi launch to reflect technology that emerged between the NGT in May 2025 and the launch in April 2026. Scenarios spanned AI-supported imaging interpretation, sepsis and deterioration prediction, ambient documentation, patient-facing AI, triage and operational analytics, and ED deployment of large language models.

### Phase 1: Theme generation (Nominal Group Technique)

The NGT session was convened on May 14, 2025, during the SAEM Annual Meeting and was co-facilitated by an EM clinical informatics faculty member and an EM resident with prior training in computer science and informatics. Six panelists were purposely recruited to represent clinical informatics and EM education expertise across geographically distributed academic institutions, including 5 EM faculty and 1 EM resident.

The session combined 2 structured activities: a post-it note exercise in which panelists individually generated and then collectively organized candidate competency statements in response to the 14 scenarios, followed by a moderated roundtable discussion in which each panelist articulated the knowledge, skills, and attitudes they considered essential to safe and effective engagement with each scenario and the group organized the themes that had emerged. The session was audio-recorded and transcribed, and the post-it artifacts were preserved. Two analysts independently coded the transcripts and post-it outputs using Braun and Clarke’s thematic analysis approach, with iterative discussion to resolve coding discrepancies.^23^ Preliminary themes were returned to the NGT panel for member review and refinement. The final inductive synthesis produced 4 themes and 10 subthemes alongside 7 candidate gap themes flagged for further development; together, these served as the items for Delphi consensus evaluation.

### Phase 2: Modified Delphi

We assembled a Delphi panel of 13 members through the working group of the 2026 SAEM AI Consensus Conference, which included 11 academic EM faculty, 1 EM resident, and 1 postdoctoral research fellow in clinical informatics, distributed across 12 academic medical centers in the West, Midwest, South, and Northeast regions of the United States. Self-reported familiarity with AI tools in clinical practice (on a five-point scale, with 1 indicating no familiarity and 5 indicating routine use) had a median of 4 (range 2–5). Among the 11 faculty panelists, years since completion of residency training had a median of 8 (range 4–28). Three panelists had also participated in the antecedent NGT session, including one of the working group co-leads; this overlap is acknowledged transparently as part of the audit trail and was addressed by structuring the Delphi panel so that most members had not contributed to the NGT.

#### Round 1

Round 1 was administered as a web-based Google Forms questionnaire that took approximately 25–30 minutes to complete; responses were collected individually and aggregated without round-level attribution to specific panelists. Structured as a divergent phase, it asked panelists to rate the face validity and clinical relevance of each of the 15 stimulus scenarios and the importance and clarity of each of the 4 themes, 10 subthemes, and 7 candidate gap themes on five-point Likert scales. Open-text prompts invited additional scenarios, themes, subthemes, or gaps not captured by the NGT. Round 1 was open from April 25 to May 2, 2026, and yielded 10 of 13 completed responses (77%).

#### Consolidation meeting

A 60-minute videoconference was convened by the working group co-leads on May 7, 2026, with 12 working group members in attendance, to deliberate on Round 1 outputs in advance of Round 2. The meeting used a tiered decision framework that distinguished items already at consensus, items requiring discussion before re-rating (the principal subject of the meeting), and items requiring fundamental reconsideration. Six scenarios were dropped as out of scope or insufficiently AI-specific (each is enumerated, with rationale, in Methods S1), 2 were combined, 1 reframed, and 1 reworded, and 2 new scenarios were added, including 1 to operationalize the post-hoc AI integration model surfaced in Round 1 free-text responses. The 4 NGT themes were retained, and a fifth theme, Cognitive impacts of AI, was added to consolidate 2 candidate gap themes (cognitive autonomy and alert fatigue) that the panel had rated as the strongest signals not captured by the original 4. The remaining gap themes were folded into existing themes as new subthemes (patient-facing AI into Theme 1; systems-level routing and interprofessional team-based AI into Theme 3) or dropped. The co-leads also proposed a one-to-one mapping from each theme to a derived competency, resulting in the expanded framework, ready for Round 2 consensus evaluation.

#### Round 2

Round 2 was administered as a web-based Google Forms questionnaire that took approximately 15–20 minutes to complete. The instrument returned to panelists the aggregated distribution of Round 1 ratings together with collated qualitative feedback for items not yet at consensus, and added items evaluating panel consensus on the architectural decisions made at the consolidation meeting (the one-to-one theme-to-competency mapping, the Theme 4 restructuring into risk-category subthemes, and the Theme 5 name). Thirty-six required items were rated, and 16 optional free-text items collected refinement suggestions. Round 2 was open from May 11 to May 15, 2026, and yielded 13 of 13 completed responses (100%).

### Post-Round 2 refinement

Following Round 2, the working group co-leads made a small number of editorial refinements to the framework based on qualitative feedback from panelists that did not require re-rating. These were limited to clarification of language and scope rather than addition or removal of framework elements, comprising 5 changes: rewording of one subtheme to bound its required depth of technical understanding; extension of another subtheme’s scope to include physician–advanced practice provider and physician–learner interactions; strengthening of a third subtheme’s descriptor to surface both deskilling and “neverskilling” concerns; narrowing of one scenario’s clinical context to acute presentations; and reframing of one scenario as a future-state operational model. Each refinement is documented in the supplemental audit trail (Table S5).

### Consensus thresholds and reporting

A priori consensus thresholds were defined as follows: scenario importance required ≥70% of panelists rating ≥4 on the five-point Likert scale with IQR ≤1; theme adequacy required ≥75% rating adequate; derived competency and subtheme adequacy each required ≥70% rating ≥4 with IQR ≤1; architectural decisions were resolved by plurality endorsement.^20,21^ Complete rating distributions and retention decisions (Table S2), architectural-decision results (Table S3), qualitative feedback and its dispositions (Methods S1), and the full decision audit trail (Table S5) are provided as supplemental material per CREDES recommendations.^21^

## THE FRAMEWORK

The consensus process produced a framework with 5 themes, 19 subthemes, 5 derived competencies, and 10 retained clinical scenarios (Figure 2, Table 1). The framework architecture rests on a one-to-one mapping from each theme to its corresponding derived competency, endorsed in Round 2 by 12 of 13 panelists as the appropriate structural choice. All 5 themes met the pre-specified threshold for adequacy (mean adequacy ratings 3.91–4.55); all 5 derived competencies met thresholds for adequacy (means 4.09–4.45); subthemes individually rated in Round 2 met consensus thresholds (rated means 4.09–4.45), while unchanged subthemes were retained based on their parent themes’ adequacy ratings; 9 of 10 scenarios met the consensus threshold for importance, with 1 retained as an explicit future-state operational model. The 5 themes trace an arc through the emergency physician’s relationship with AI: how physicians communicate about it (Theme 1), judge when to use it (Theme 2), work with it in real time (Theme 3), manage what it gets wrong (Theme 4), and protect their own reasoning alongside it (Theme 5). Each theme is presented below with its derived competency and subthemes; complete consensus statistics for every rated element appear in Table 3.

**Table 1.**
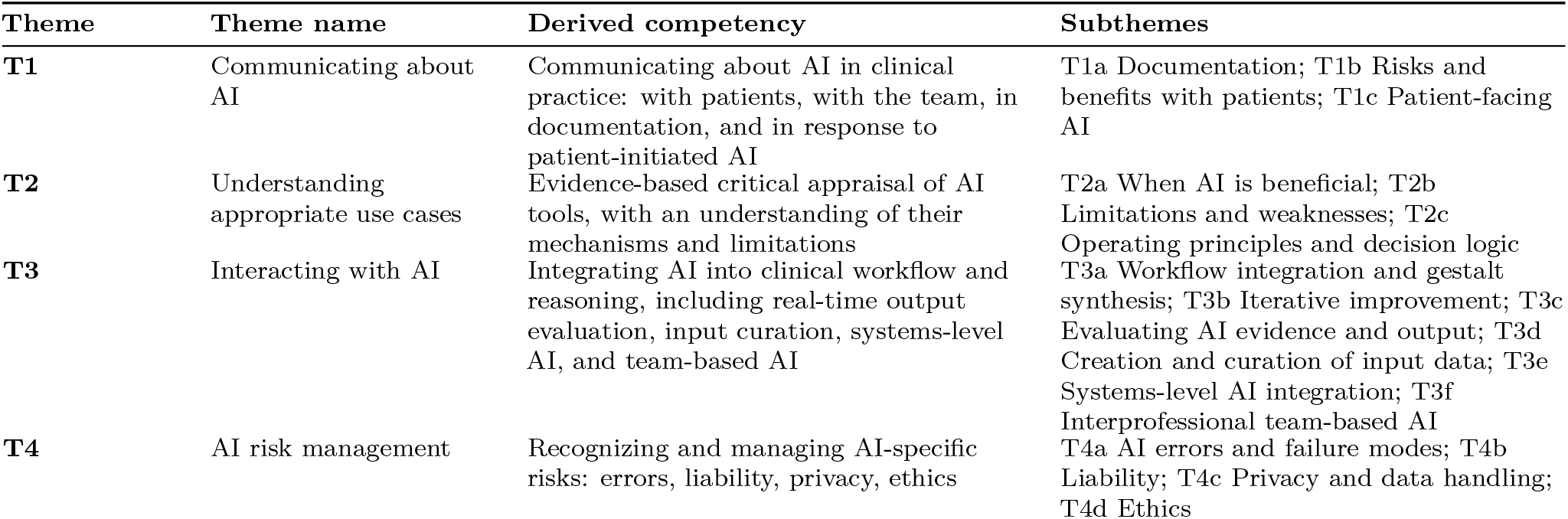
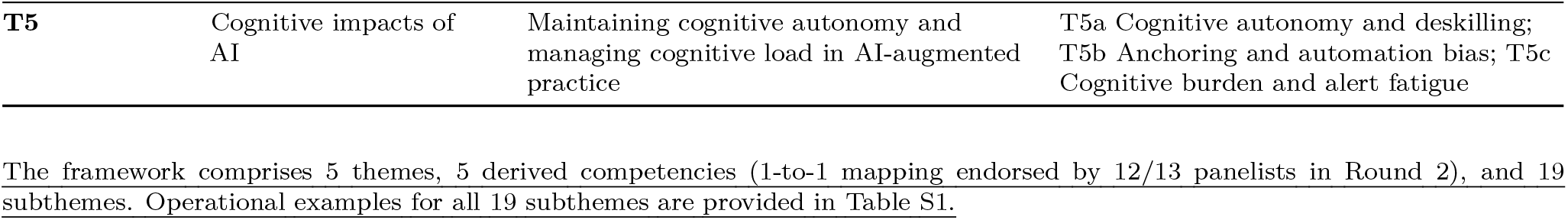
Framework structure: five themes, derived competencies, and subthemes.

**Table 2.** Ten retained clinical scenarios.

| Id | Title | Primary themes surfaced | Round 1 importance | Round 2 importance | Status |
| --- | --- | --- | --- | --- | --- |
| S1 | AI history-of-present-illness summarization | T1a, T2b, T3c | 4.00 · 75% · IQR 1.0 | Not re-rated | Passed R1 |
| S4 | AI ECG interpretation flagging Brugada pattern | T2b, T3a, T3c, T1c | 4.50 · 100% · IQR 0.0 | Not re-rated | Passed R1 |
| S6 | AI stroke imaging localization | T2b, T3a, T3c | 4.00 · 75% · IQR 1.0 | Not re-rated | Passed R1 |
| S9 | Smartwatch palpitations | T1c, T2a, T3c | 3.88 · 75% · IQR 1.0 | Not re-rated | Passed R1 |
| S10 | AI image interpretation | T2a, T2b, T3c, T5b | 3.62 · 62% · IQR 1.0 | 4.09 · 82% · IQR 1.0 | Passed R2 |
| S2+S15 | AI-assisted documentation (ambient and inline) | T1a, T3a, T3c, T4a | S15: 4.62 · 88%; S2: 3.12 · 50% | 4.27 · 91% · IQR 1.0 | Passed R2 (combined) |
| S11 | Pre-emptive AI consultation | T3a, T3c, T5a, T5b | 4.00 · 50% · IQR 2.0 | 4.18 · 91% · IQR 1.0 | Passed R2 (reframed) |
| S14 | AI resource utilization and patient flow | T3e, T3f, T2b | 3.62 · 62% · IQR 2.2 | 4.27 · 91% · IQR 1.0 | Passed R2 (reworded) |
| S16 | Patient-initiated AI consultation | T1b, T1c, T2a | New in R2 | 4.36 · 91% · IQR 1.0 | Passed R2 (new) |
| S17 | Post-hoc AI review | T3c, T5a | New in R2 | 3.82 · 55% · IQR 1.5 | Borderline (retained as future-state) |

Round 1 statistics for scenarios passing in R1 are shown for reference; those scenarios were not re-rated in R2 and were retained on confirmation. Importance reported as mean on a 5-point Likert scale · proportion of panel rating $\geq 4$ · interquartile range. Round 1 panel n = 10; Round 2 panel n = 13. S15 = ambient AI scribe documentation; S2 = inline autocompletion.

**Table 3.**
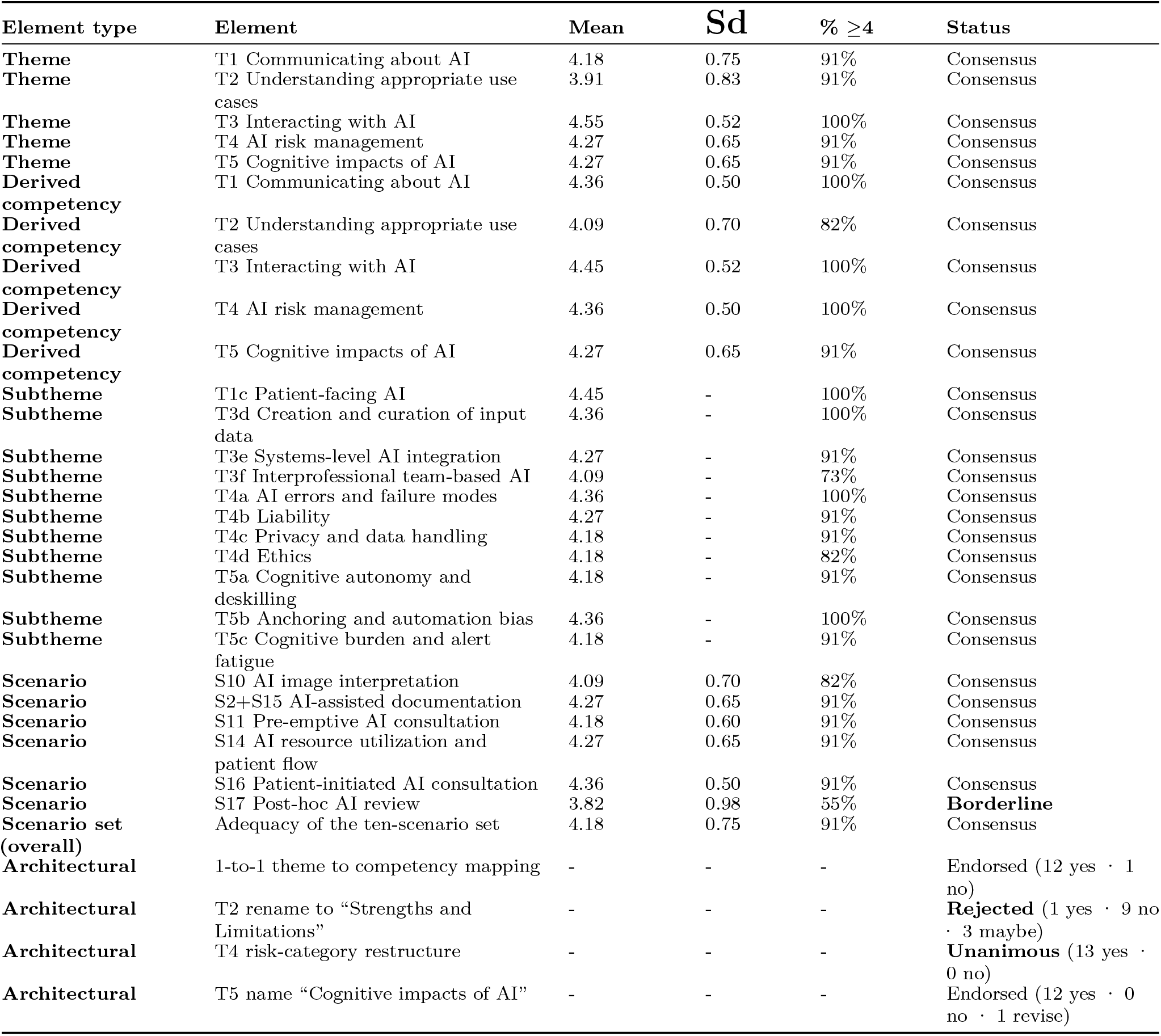
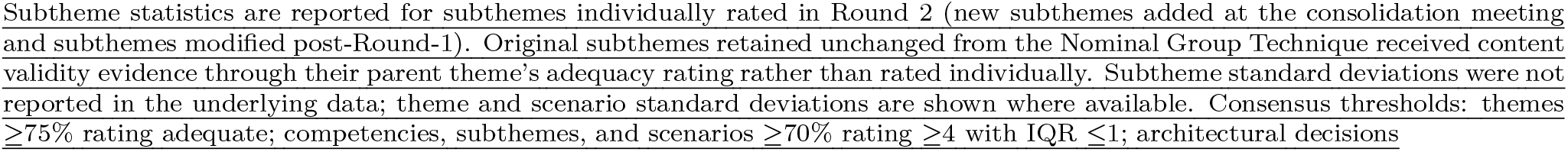
Consensus statistics for all rated framework elements (Round 2, n = 13)

**Figure 2.**
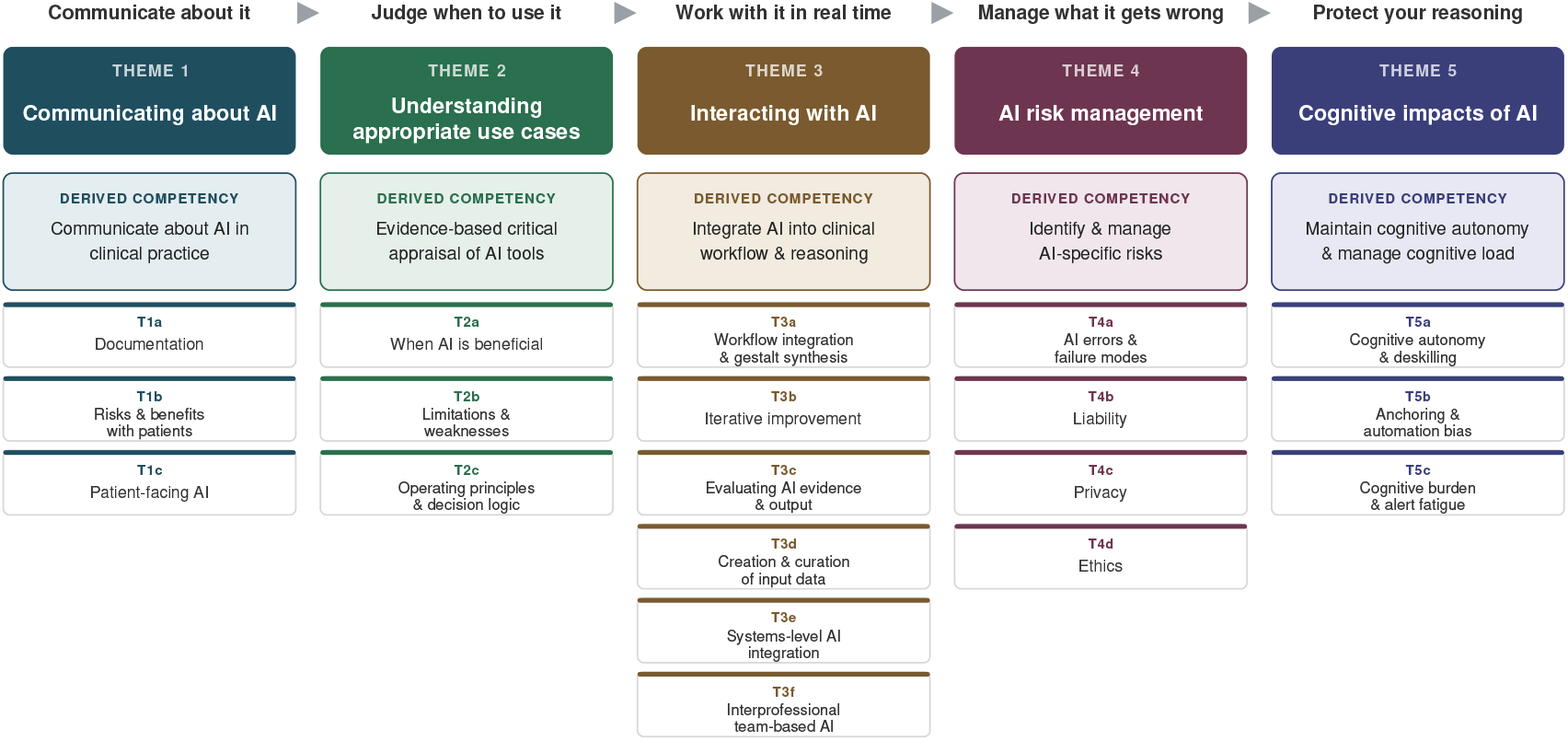
A consensus competency framework for physician use of artificial intelligence in emergency medicine. Five themes trace the emergency physician’s relationship with AI across the clinical encounter: communicating about AI, understanding appropriate use cases, interacting with AI, managing AI-specific risks, and the cognitive impacts of AI. Each theme carries a single derived competency (shaded) and its constituent subthemes (T1a–T5c). AI, artificial intelligence.

### Theme 1: Communicating about AI

Theme 1 encompasses the verbal and written communication competencies emergency physicians must develop to operate within an AI-augmented clinical environment: communication with patients about AI’s role in their care, with the clinical team about AI-derived information, through documentation where AI-generated content increasingly appears, and in response to patient-initiated AI use. The derived competency is the ability to communicate about AI in clinical practice across these four contexts. Three subthemes operationalize this scope: <u>T1a Documentation</u>, addressing the disclosure, attestation, and reconciliation of AI-generated content within the medical record; <u>T1b Risks and benefits with patients</u>, addressing physician-led communication about AI-supported clinical decisions and informed consent; and <u>T1c Patient-facing AI</u>, addressing physician response to patient-initiated AI during the clinical encounter, including patients arriving with AI-generated differentials, AI-summarized chart access, and AI interpretations of patient-generated wearable data. All Theme 1 elements met consensus thresholds; T1c received the strongest endorsement of any new subtheme in Round 2 (mean 4.45, 100% rated ≥4), reflecting a consistent qualitative signal from Round 1 that identified patient-facing AI as the most prominent gap in the original four-theme framework.

### Theme 2: Understanding appropriate use cases

Theme 2 addresses physicians’ understanding of when AI tools are useful, their limitations, and what operational understanding of their mechanisms is required to use them appropriately. AI evidence has features that distinguish it from other forms of clinical evidence (training data provenance, scale, opacity, hallucination tendencies, and distribution shift) that require updated appraisal skills. The derived competency is evidence-based critical appraisal of AI tools, with an understanding of their mechanisms and limitations. Three subthemes operationalize this scope: <u>T2a When AI is beneficial</u>, identifying clinical contexts in which AI tools provide value and recognizing when not to use them; <u>T2b Limitations and weaknesses</u>, recognizing AI-specific failure modes (hallucination, trainingdata drift, distribution shift, subgroup performance disparity) and the conditions under which output should be discounted; and <u>T2c Operating principles and decision logic</u>, understanding how AI tools generate their output to a degree sufficient to evaluate it critically, explicitly bounded to exclude any requirement for machine-learning technical expertise. Theme 2 received the lowest theme adequacy in the framework (mean 3.91, 91% rated ≥4), reflecting substantive Round 2 debate about the appropriate depth of mechanistic understanding emergency physicians need; the T2c rewording was the post-Round-2 refinement most directly responding to that debate.

### Theme 3: Interacting with AI

Theme 3 addresses how emergency physicians use AI tools in clinical practice, including workflow integration, iterative improvement, real-time output evaluation, input curation, systems-level AI operating upstream of physician interaction, and AI’s effects on interprofessional team dynamics. The derived competency is integrating AI into clinical workflow and reasoning, including real-time output evaluation, input curation, systems-level AI, and team-based AI. Six subthemes operationalize this scope, reflecting the breadth of physician–AI interaction modes. <u>T3a Workflow integration and gestalt synthesis</u> addresses integrating AI-derived information into clinical reasoning without disrupting workflow. <u>T3b Iterative improvement</u> addresses adjusting AI use over time based on observed performance and contributing to local quality improvement processes. <u>T3c Evaluating AI evidence and output</u> addresses real-time critical appraisal of AI-generated content; the in-the-moment evaluation of a specific output, operating downstream of <u>T2c. T3d Creation and curation of input data</u> addresses composing prompts, structuring inputs, and curating the information given to AI tools to optimize output quality. <u>T3e Systems-level AI integration</u> addresses AI systems performing operational functions in the ED (patient flow routing, triage scoring, lab ordering from triage data, resource allocation, disposition prediction) that operate upstream of physician evaluation. <u>T3f Interprofessional team-based AI</u> addresses AI’s effect on interprofessional team interactions (physician– nurse, physician–advanced practice provider, physician–learner, physician–technologist, physician–EMS) and AI’s use as a mediator in these interactions; the post-Round-2 scope extension to physician–advanced practice provider and physician–learner interactions responded to specific panel feedback. Theme 3 received the strongest theme adequacy in the framework (mean 4.55, 100% rated ≥4); the derived competency was endorsed by 100%.

### Theme 4: AI risk management

Theme 4 addresses the categories of risk that emergency physicians must recognize and manage when AI tools contribute to clinical decisions. The theme was renamed during the consolidation meeting from the original NGT framing, “Medico-legal risk and governance,” to reflect the principle that the core competency is recognizing what AI gets wrong and the consequences when it does; recognizing that liability, regulatory, and governance frameworks are the operational context within which competency is exercised, not the competency itself. The derived competency is identifying and managing AI-specific risks across errors, liability, privacy, and ethics. Four subthemes structure this scope: <u>T4a AI errors and failure modes</u> addresses recognizing and managing AI-specific failure modes (hallucinations, bias, drift, distribution shift, sycophancy), operating downstream of T2b with focus on response and management; <u>T4b Liability</u> addresses allocation and management of legal exposure when AI contributes to clinical decisions or errors; <u>T4c Privacy and data handling</u> addresses Health Insurance Portability and Accountability Act (HIPAA) and Family Educational Rights and Privacy Act (FERPA) implications, secondary use of clinical data, and patient consent for AI processing; and T4d Ethics addresses informed consent for AI involvement, equity considerations including subgroup performance and access disparities, patient autonomy, and just institutional deployment. All Theme 4 elements met consensus thresholds, and the risk-category restructuring was unanimously endorsed by all 13 panelists in Round 2 as preferable to the original regulatory and governance scaffolding.

### Theme 5: Cognitive impacts of AI

Theme 5 addresses the cognitive consequences of working in an AI-augmented practice environment, both at the level of individual reasoning (maintaining clinical judgment when AI offers recommendations) and at the level of cognitive load (managing AI-generated alerts and recommendations alongside the existing clinical decision-support burden). The theme was added during the consolidation meeting by combining two candidate gap themes (cognitive autonomy and alert fatigue) that the Round 1 panel rated as the strongest standalone signals not captured by the original four themes. The derived competency is maintaining cognitive autonomy and managing cognitive load in AI-augmented practice. Three subthemes structure this scope: T5a Cognitive autonomy and deskilling addresses maintaining independent clinical reasoning when AI tools surface recommendations, explicitly naming both deskilling (degradation of established competence in practicing physicians) and “neverskilling” (failure to develop competence in trainees who rely on AI before building baseline skills);^24,25^ T5b Anchoring and automation bias addresses resisting the cognitive pull of AI-suggested differentials, disposition decisions, or risk scores when clinical judgment disagrees;^26^ and T5c Cognitive burden and alert fatigue addresses managing the cognitive load of AI-generated alerts and the recognition that alert fatigue itself can become a patient safety risk.^27^ All Theme 5 elements met consensus thresholds; the theme name “Cognitive impacts of AI” was endorsed by 12 of 13 panelists.

### Clinical scenarios

The framework’s 10 retained clinical scenarios (Table 2; full vignettes are provided in Table S4) anchor the competency themes in concrete clinical situations; they are intended to span the AI competency landscape sufficiently for educational use rather than to enumerate every encounter emergency physicians may face. Four scenarios passed Round 1 consensus without modification (AI history-of-present-illness summarization, AI electrocardiogram [ECG] interpretation, AI stroke imaging, and smartwatch palpitations); 5 additional scenarios reached consensus in Round 2 after consolidation-meeting refinement (AI image interpretation, combined AI-assisted documentation, pre-emptive AI consultation, AI resource utilization and patient flow, and patient-initiated AI consultation). One scenario (post-hoc AI review) remained borderline at Round 2 (mean 3.82, 55% rated ≥ 4) but was retained as an explicit future-state operational model, reflecting the conceptual importance of the pre-emptive versus post-hoc integration axis discussed below. The overall scenario set adequacy was endorsed by 91% of the panel (mean 4.18).

### Architectural decisions

Four architectural decisions were submitted to Round 2 for consensus evaluation, along with the theme and subtheme ratings. The one-to-one mapping from each theme to a derived competency was endorsed by 12 of 13 panelists as the appropriate structural choice, treating themes as primary and competencies as a derived layer consistent with established medical education competency frameworks. A proposed renaming of Theme 2 from “Understanding appropriate use cases” to “Strengths and Limitations” was rejected by the panel (9 no, 3 maybe, 1 yes); the original name was retained. The risk-category restructuring of Theme 4 into Errors, Liability, Privacy, and Ethics, replacing the original regulatory and governance scaffolding, was unanimously endorsed by all 13 panelists. The name “Cognitive impacts of AI” for Theme 5 was endorsed by 12 of 13, with one panelist suggesting “Cognitive challenges of AI” as an alternative; the original name was retained. Complete consensus statistics for every rated framework element are provided in Table 3.

### Audit trail

The framework’s full decision provenance from NGT through post-Round-2 refinement is documented in Table S5 and Methods S1. The most substantive evolution from NGT output to final framework occurred at the consolidation meeting, as detailed in the Methods. All framework elements reached consensus in Round 2; the post-Round-2 refinement applied five editorial changes without altering the underlying structure.

## DISCUSSION

The present framework addresses a gap that persists across medical education—graduate requirements omit AI and the AAMC’s forthcoming undergraduate competencies are deliberately specialty-agnostic—through specialty-specific anchoring in 10 clinical scenarios, quantitative content-validity evidence,^28^ and explicit elevation of cognitive autonomy and alert fatigue, domains not surfaced in prior generalist models.

### Pre-emptive versus post-hoc AI integration

A conceptual axis emerged from the consensus process that cuts across the framework and has direct implications for educational design. AI tools can be integrated into clinical workflow in two fundamentally different ways. In the pre-emptive model, AI surfaces recommendations, risk scores, or differential diagnoses before the clinician completes independent reasoning; examples include active consultation tools, pop-up clinical decision support such as sepsis alerts,^3^ and AI-generated triage scores.^1^ In the post-hoc model, AI reviews the physician’s completed work and flags potential omissions or errors; examples include post-encounter chart review tools and AI that reads a completed note to suggest broader differentials. The framework’s paired scenarios S11 (pre-emptive consultation) and S17 (post-hoc review) deliberately surface the same physician–AI interaction problem under both models.

The two integration modes have distinct cognitive implications. The pre-emptive model produces well-documented risks of anchoring and deskilling because the clinician’s reasoning is shaped by AI input before independent thinking is complete; for trainees, this raises the additional concern of neverskilling, in which foundational reasoning skills fail to develop because they were never practiced under conditions that required them.^34^ The post-hoc model preserves the development of independent clinical reasoning; AI serves as a safety net rather than a substitute. A recent study of clinicians using a post-hoc AI consultation tool in primary care documented a co-learning effect, with independent clinical performance improving over time alongside continued AI exposure.^29^ The framework does not endorse one integration model over the other but prepares physicians to recognize and operate within both, with the cognitive autonomy and anchoring competencies in Theme 5 applying differently in each case.

### Tiered competencies as articulated need

Round 1 showed strong support for differentiating competencies at the medical-student level (7 of 10 panelists endorsed substantial differentiation), with weaker signals up the training continuum. The consolidation meeting resolved that the framework would articulate the need for tiered competencies but not pre-specify which competencies attach to which stage. This positioning reflects both methodological and empirical considerations. Methodologically, stage-specific assignment is a curriculum-development task requiring local context (program structure, patient population, institutional AI deployment) and consensus among educators within a particular training stage rather than within an AI competency working group; a framework that pre-assigned competencies to stages would be either too rigid for diverse contexts or too generic to be useful. Empirically, recent work mapping EM resident clinical exposure against the ABEM Model of Clinical Practice has shown wide individual variability: graduating residents have encountered between 59.4% (532/895) and 67.3% (602/895) unique clinical topics with substantial overlap across postgraduate years,^13^ a pattern that undermines the assumption that competency progression maps cleanly to training year. Stage-specific competency assignment is captured as an explicit research priority in the conference’s broader research agenda.

### Position relative to existing competency frameworks

The most directly comparable prior work is the six-domain framework for AI competencies in health care professionals introduced above, derived from qualitative interviews with 15 subject-matter experts.^18^ That framework’s domains (basic knowledge of AI, social and ethical implications, workflow analysis, AI-enhanced clinical encounters, evidence-based evaluation, and practice-based learning) overlap substantially with the framework presented here, particularly in evidence-based evaluation (Theme 2) and AI-enhanced clinical encounters (Themes 1 and 3). Three differences are worth noting. First, the prior framework is profession-wide by design, intended to define a baseline applicable to all health care professionals, whereas ours is specialty-specific and anchored in EM scenarios. Second, the prior framework, while qualitatively sound, was generated without quantitative consensus evaluation; ours has content validity evidence from quantitative panel consensus against pre-specified thresholds and documented architectural endorsement. Third, while the prior framework acknowledges anchoring and automation bias, ours operationalizes those concerns for the EM context and extends them to include alert fatigue and cognitive load. The 2 frameworks are thus best understood as complementary: a profession-wide baseline remains useful, and ours translates that baseline into specialty-specific terms.

The AAMC’s parallel cross-continuum Delphi process targets a fall 2026 release;^17^ the framework presented here is consistent with its competency-based approach and could provide an operational template for specialty-specific extensions when the national framework is released. Internationally, the DECODE consortium used a modified Delphi process to develop digital health competencies for medical students across multiple countries,^22^ with results emphasizing cross-cutting digital health literacy rather than AI specifically; this provides methodological precedent for the design choice here, though the scope is broader and less AI-focused.

### Implications for training, assessment, and faculty development

The framework provides a structural target for EM training programs working to integrate AI competencies into existing curricula. For undergraduate medical education, the themes can be mapped to existing curricular touchpoints (clinical reasoning, evidence-based medicine, professionalism, communication, and informatics), allowing AI-specific content to be added without standalone curriculum development. For graduate medical education, the competencies are candidates for inclusion in residency milestone development; the absence of AI from current ACGME Program Requirements^15^ makes such inclusion both feasible and timely. For continuing medical education, the framework defines a content scope for faculty development modules, a particular concern given that practicing emergency physicians vary widely in their exposure to AI tools, and Theme 5 explicitly addresses the deskilling concern affecting this cohort.

Assessment instruments with validity evidence supporting their use for AI competencies do not yet exist in any specialty. The framework defines the targets that assessment instruments would measure, but does not prescribe methodology. Workplace-based assessment (for example, observing how a resident integrates an AI-flagged finding into clinical reasoning or how a physician reviews ambient scribe output) is a natural extension of established entrustable professional activity frameworks. A related strand of graduate medical education work has begun to address how AI tools should be incorporated into supervision and entrustment decisions,^24,30–33^ providing practical guidance for program directors that complements the framework presented here.

## LIMITATIONS

Several limitations should inform the interpretation and application of the framework. The Delphi panel of 13, while consistent with established methodological norms, is small; additional content validity evidence from emergency physicians outside academic medical centers, in community-practice settings, and internationally will strengthen external validity. Three of the 13 Delphi panelists also participated in the antecedent NGT session, disclosed in the audit trail and structurally addressed by recruiting the majority of the Delphi panel independent of the NGT, though it remains a source of non-independence. Theme 2’s lower adequacy rating reflects unresolved debate about the depth of mechanistic understanding emergency physicians need; the post-Round-2 refinement of T2c clarifies the framework’s position but does not eliminate the underlying tension. The framework is current with the 2026 AI deployment landscape; rapid evolution, particularly in large language models, ambient documentation, and patient-facing AI, will require periodic re-evaluation on an anticipated 18- to 24-month cycle. Finally, the framework articulates competencies but does not assess them; inter-rater reliability of the operational examples (Table S1) among educators has not been established.

## FUTURE DIRECTIONS

Several lines of work follow. Curriculum operationalization is the most immediate, with stage-specific competency mapping (medical student, intern, junior resident, senior resident, attending) as the central downstream task. Assessment instruments with validity evidence supporting their use for each derived competency are needed and constitute an explicit priority in the conference’s broader research agenda. Re-evaluation through a subsequent consensus process in 18 to 24 months will allow the framework to incorporate the next generation of AI tools, particularly in cognitive impacts and patient-facing AI, where current deployment is least mature. Two scope additions are anticipated: a framework addendum on AI in emergency medical services and competencies for emerging genomic and omics-based AI applications as these technologies move closer to EM clinical relevance. Integration with the forthcoming AAMC AI competencies will allow this framework to function as a specialty-specific operationalization of the national cross-continuum baseline.

## CONCLUSION

We present an EM-specific framework for AI competency comprising five themes, 19 subthemes, five derived competencies, and 10 retained clinical scenarios. To our knowledge, this is the first specialty-specific consensus AI competency framework for a United States medical specialty and the first to elevate cognitive autonomy and alert fatigue to distinct AI competency subdomains. Its ultimate value will depend on uptake by residency programs, medical schools, and continuing medical education providers, and on whether physicians trained using this framework can integrate AI into clinical care while maintaining independent clinical judgment, as the AI deployment landscape continues to evolve.

### Generative AI Disclosure

The authors used generative AI tools (including OpenAI ChatGPT, Anthropic Claude, and Google Gemini) to varying degrees at several stages of the research and writing process, including brainstorming and refining ideas, obtaining critical feedback on analytic and conceptual approaches, and reviewing and editing language for clarity and concision. All AI-assisted output was critically reviewed, verified, and edited by the authors. The intellectual content, analyses, interpretations, and conclusions of this manuscript represent the authors’ own work, and the authors take full responsibility for the integrity and accuracy of the manuscript.

## Data Availability

The data that support the findings of this study are available in the supplementary material accompanying this preprint. Further data are available from the corresponding author upon reasonable request.

