## Supplementary Material for "An AI Competency Framework for Emergency Medicine: A Multiphase Consensus Process"

### Supplementary Appendix

#### Contents

Methods S1. Expanded methods, dropped-scenario rationale, and Round 2 qualitative dispositions
Table S1. Operational examples for all 19 subthemes
Table S2. Round 2 consensus statistics for all rated elements
Table S3. Architectural validation results (Round 2)
Table S4. The ten clinical scenarios: full vignettes and consensus status
Table S5. Decision audit trail (nominal group technique through version 3)

#### Methods S1. Expanded methods, dropped-scenario rationale, and Round 2 qualitative dispositions

##### Consensus design

The framework was developed through a sequential nominal group technique (NGT)-to-Delphi design. Phase 1 used an NGT to inductively generate candidate themes and subthemes from a curated set of clinical AI scenarios; Phase 2 used a two-round modified Delphi process, separated by a virtual consolidation meeting, to evaluate and refine the resulting architecture, followed by a post-Round 2 refinement phase in which the working group co-leads applied editorial revisions based on panel qualitative feedback. Reporting of the Delphi component followed the Guidance on Conducting and REporting DElphi Studies (CREDES).

##### Consensus thresholds (a priori)

Scenario importance required at least 70% of panelists rating the item 4 or higher on a 5-point Likert scale with an interquartile range (IQR) $\leq 1$. Theme adequacy required at least 75% of panelists rating the theme adequate. Derived-competency and subtheme adequacy each required at least 70% rating $\geq 4$ with IQR $\leq 1$. Architectural decisions were resolved by plurality endorsement. Elements meeting one threshold but not the other were designated borderline and retained only with explicit justification.

##### Panel composition and independence

The Round 2 Delphi panel comprised 13 members distributed across 12 academic medical centers. Three of the 13 panelists had also participated in the antecedent nominal group session, including one of the working group co-leads. This overlap is disclosed for transparency; it was structurally mitigated by recruiting the majority of the panel from members who had not contributed to the NGT, though it remains a source of non-independence.

##### Dropped scenarios

Six Round 1 scenarios were dropped during the May 7, 2026 consolidation meeting. They are enumerated here with rationale for methodological transparency.

- **S3 Insurance billing AI -** too institution-specific and policy-dependent.
- **S5 Transcriptome pulmonary-embolism risk -** genomics not anticipated to enter emergency medicine within the relevant timeframe; noted for a future framework addendum.
- **S7 Delirium-risk pop-up -** not novel; existing alert-fatigue literature already addresses the underlying competency.
- **S8 Paracentesis harm prediction -** vague and not AI-specific to emergency medicine.
- **S12 Mortality-prediction alert -** functionally a goals-of-care competency rather than an AI competency.
- **S13 Prehospital surgical prediction -** EMS-specific; outside the working group's residency-training scope.

##### Scenario proposed in Round 2 but not included as a standalone vignette

One Round 2 panelist proposed a medicolegal failure scenario: a chart-summarizing AI hallucinates a severe contrast allergy the patient never had; the stroke team withholds CT angiography on that basis; definitive diagnosis of a large ischemic stroke is delayed; and the family pursues litigation over the resulting harm. The version-3 decision is to use this as an illustrative vignette within the Theme 4 section of the concept paper rather than as a standalone framework scenario, because it is already covered by subtheme T4a (AI errors and failure modes) as operationalized through S1 (summarization with hallucinated content) and S4 (a flagged finding requiring physician adjudication).

##### Round 2 qualitative themes and version-3 dispositions

Major qualitative signals from Round 2 free-text responses, each with the disposition applied in version 3.

**T2c "under-the-hood mechanisms" critique.** A panelist objected (twice) that Theme 2 risked scope creep and that emergency physicians do not need to understand AI tool internals to use them appropriately. Disposition: the subtheme was reworded to "Operating principles and decision logic," and the concept paper defends the operational-versus-technical distinction explicitly.

**S17 borderline status.** A panelist summarized the panel view that post-hoc review is probably the future but may not be the highest priority. Disposition: S17 retained with explicit future-state framing, because the pre-emptive versus post-hoc axis matters to the framework even where S17 is currently aspirational.

**S2+S15 combination contested.** Two panelists argued that ambient scribes and autocompletion tap different cognitive risks and should be split. Disposition: the combined scenario was retained for parsimony (91% endorsement), and the concept paper acknowledges the two distinct cognitive-risk profiles.

**T3f scope extension.** A panelist requested explicit inclusion of physician–advanced practice provider and physician–learner interactions. Disposition: the T3f definition and examples were extended to name both.

**T5a deskilling/neverskilling language.** A panelist noted the descriptor did not adequately surface the deskilling concern. Disposition: the T5a definition was strengthened to name both deskilling (practicing physicians) and neverskilling (trainees).

**S16 refinements.** A panelist proposed narrowing S16 to an acute presentation, moving wearables to S9, and changing "ChatGPT" to "chatbot." Disposition: all three refinements were applied.

##### Table S1. Operational examples for all 19 subthemes

Operational examples illustrate each subtheme; they are not assessment items, and inter-rater reliability among educators has not been established. Subthemes are grouped under their parent theme, which maps one-to-one to a single derived competency.

| Subtheme | Definition and operational examples |
|---|---|
| **Theme 1 · Communicating about AI** | |
| **T1a**<br>Documentation | **Definition.** Communicating AI-generated content within the medical record, including disclosure of AI involvement, attestation of accuracy by the physician, and management of the discrepancies that arise between AI-generated text and physician judgment.<br>Operational examples:<br>• Reviewing and editing an ambient AI scribe note before signing, attesting that the documented content accurately reflects the encounter<br>• Documenting that an AI-flagged finding (e.g., EKG Brugada pattern) was reviewed and adjudicated by the physician<br>• Disclosing AI assistance in clinical documentation per institutional and regulatory requirements |

| Subtheme | Definition and operational examples |
|---|---|
| **T1b**<br>Risks and benefits with patients | **Definition.** Physician-led communication with patients about AI-supported clinical decisions, including informed consent for AI-involved care, explanation of AI's role in diagnostic or therapeutic recommendations, and management of patient uncertainty about AI's reliability.<br>Operational examples:<br>• Explaining to a patient that an AI tool flagged an imaging abnormality and that the physician has reviewed and adjudicated the finding<br>• Discussing with a patient when an AI-recommended workup may add value versus when it can be safely deferred<br>• Addressing patient questions about whether AI is replacing physician judgment |
| **T1c**<br>Patient-facing AI | **Definition.** Physician response to patient-initiated AI use during the clinical encounter, including patients arriving with AI-generated differentials from chatbots, AI-summarized chart access via patient portals, AI-generated patient education they believe is authoritative, and patient-generated wearable data with AI interpretations (smartwatch arrhythmia detections, continuous glucose monitor trends, sleep-tracking AI). T1c absorbs the former gap theme G2 (patient-generated data and wearables) since wearable data interpretation increasingly relies on AI and surfaces to patients before clinicians.<br>Operational examples:<br>• Responding to a patient who arrives with a ChatGPT-generated differential and requests a specific workup<br>• Integrating smartwatch-flagged arrhythmia data into clinical reasoning and patient communication<br>• Managing the AI-summarized version of the patient's chart that the patient accessed via the patient portal before evaluation<br>• Discussing AI-generated patient education that may contain inaccuracies relative to current evidence |
| **Theme 2 · Understanding appropriate use cases** | |
| **T2a**<br>When AI is beneficial | **Definition.** Identifying clinical contexts in which AI tools provide diagnostic, therapeutic, operational, or educational value. Includes the corresponding ability to recognize when NOT to use a tool: when patient context, data quality, or workflow conditions render an AI tool unreliable or low-value.<br>Operational examples:<br>• Identifying that AI image interpretation adds value for screening abnormalities on CXR but should not substitute for physician interpretation in high-stakes calls<br>• Recognizing that LLM-generated discharge summaries may be appropriate for routine encounters but should be carefully reviewed for atypical cases<br>• Knowing when an AI triage tool should be overridden based on physician gestalt |

| **Subtheme** | **Definition and operational examples** |
|---|---|
| **T2b**<br>Limitations and weaknesses | **Definition.** Recognizing AI-specific failure modes (hallucination, training-data drift, distribution shift, sycophancy, subgroup performance disparity) and the conditions under which a tool's output should be discounted or overridden. This subtheme operates upstream of T4a (response and management of AI errors): T2b is awareness; T4a is institutional response.<br>Operational examples:<br>• Recognizing that an LLM-generated chart summary may contain hallucinated findings (e.g., reported allergies that the patient does not have)<br>• Identifying when an AI model's performance may degrade for patients whose presentation or demographics differ from the training distribution<br>• Understanding that AI image interpretation tools may be over- or under-sensitive in specific clinical scenarios |
| **T2c**<br>Operating principles and decision logic | **Definition.** Understanding how AI tools generate their output (what data they use, what their validation status is in the deployment context, and what their decision logic does at the operational level) to a degree sufficient to evaluate the output critically. This subtheme does NOT require machine-learning expertise; it requires operational mechanism understanding.<br>Operational examples:<br>• Knowing that an LLM-generated answer reflects pattern matching against training data rather than reasoning from first principles<br>• Understanding that an AI risk-prediction tool depends on the variables it was trained with and if those variables are missing or unreliable in the deployed setting, the prediction degrades<br>• Recognizing that an AI image interpretation tool may have been validated on different scanner protocols or patient populations than those in your ED |
| **Theme 3 · Interacting with AI** | |
| **T3a**<br>Workflow integration and gestalt synthesis | **Definition.** Integrating AI-derived information into clinical reasoning without disrupting workflow; synthesizing AI output with physical exam findings, history, team input, and clinical gestalt to form an integrated assessment.<br>Operational examples:<br>• Incorporating an AI-flagged EKG finding into the broader clinical assessment of a patient with chest pain<br>• Using AI-generated triage scores as one input among several rather than as a definitive disposition<br>• Adjusting the pace of clinical workflow when AI tools require additional review steps |
| **T3b**<br>Iterative improvement | **Definition.** Adjusting AI use over time based on observed performance, error patterns, and changing tool capabilities. Includes both individual practice adjustment and contribution to local quality improvement processes around AI deployment.<br>Operational examples:<br>• Recognizing that a particular AI tool is producing increasing false positives in your patient population and adjusting reliance<br>• Reporting an AI error or failure mode through institutional feedback channels<br>• Adapting documentation review practices as AI scribe accuracy improves over successive software updates |

| Subtheme | Definition and operational examples |
|---|---|
| **T3c**<br>Evaluating AI evidence and output | **Definition.** Real-time critical appraisal of AI-generated content for accuracy, appropriateness, and consistency with clinical context. Operates downstream of T2c (understanding how the tool works in general). T3c is the in-the-moment evaluation of a specific output.<br>Operational examples:<br>• Reviewing an LLM-generated differential diagnosis and identifying items that do not fit the clinical context<br>• Catching a hallucinated finding in an AI-generated chart summary before it influences clinical decisions<br>• Recognizing when an AI-flagged finding represents a genuine concern versus an artifact or false positive |
| **T3d**<br>Creation and curation of input data [broadened in v2] | **Definition.** Composing prompts, structuring inputs, and curating the information given to AI tools to optimize output quality. Broadened in v2 from the original NGT subtheme "prompt engineering," which was a narrower subdomain.<br>Operational examples:<br>• Crafting a clinical query for an LLM consultation tool that provides sufficient context for a useful response<br>• Selecting which portions of the clinical history are most relevant to share with an AI consultation tool<br>• Structuring an AI image interpretation request in a way that surfaces clinically relevant findings |
| **T3e**<br>Systems-level AI integration [NEW in v2; absorbs G4] | **Definition.** AI systems performing operational functions in the ED (patient flow routing, triage scoring, lab ordering from triage data, resource allocation, disposition prediction). Physician interaction with AI that performs upstream decisions affecting clinical workflow, often before the physician has evaluated the patient.<br>Operational examples:<br>• Understanding why a particular patient was brought back from the waiting room before others based on AI triage scoring<br>• Recognizing when AI-initiated lab orders from triage data align with clinical assessment versus require additional or different studies<br>• Overriding AI disposition predictions when clinical judgment disagrees |
| **T3f**<br>Interprofessional team-based AI [NEW in v2; absorbs G5; scope extended in v3] | **Definition.** AI's effect on interprofessional team interactions (physician-nurse, physician-APP, physician-learner, physician-tech, physician-EMS) and the use of AI as a mediator in these interactions. v3 explicitly extends scope to include physician-APP supervisory interactions and physician-learner teaching interactions in response to R2 panelist feedback.<br>Operational examples:<br>• Discussing AI-flagged findings with nursing colleagues during shift handoffs<br>• Supervising an advanced practice provider (APP) whose clinical workup was AI-assisted, and adjudicating the use of AI in the workup<br>• Pulling up an AI clinical reference tool during teaching with a resident to settle a disagreement about workup choices<br>• Communicating AI-derived information to EMS during prehospital handoff |
| **Theme 4 · AI Risk Management** | |

| Subtheme | Definition and operational examples |
|---|---|
| **T4a**<br>AI errors and failure modes | **Definition.** Recognizing and managing AI-specific failure modes (hallucinations, bias [in training data and in deployment], drift, distribution shift, sycophancy, brittleness on out-of-distribution inputs). Includes reporting workflows, override protocols, and documentation requirements when AI is identified as wrong. Operates downstream of T2b (recognition); T4a is the response and management dimension.<br>Operational examples:<br>• Knowing the institutional reporting workflow when an AI tool produces a clinically significant error<br>• Documenting an AI override decision in a way that protects the physician and preserves the audit trail<br>• Recognizing that an LLM is hallucinating a contrast allergy that the patient does not have, and acting to prevent that hallucination from cascading into clinical decisions |
| **T4b**<br>Liability | **Definition.** Allocation and management of legal exposure when AI contributes to clinical decisions or errors. Includes individual versus institutional responsibility, documentation practices that protect physicians while preserving accuracy, and emerging legal precedent.<br>Operational examples:<br>• Understanding the documentation requirements when an AI scribe-generated note contains errors that are signed and become part of the medical record<br>• Recognizing the liability implications of overriding versus following an AI recommendation<br>• Adapting practice to emerging litigation patterns around ambient AI use (e.g., recent cases flagged in the May 7 consolidation meeting) |
| **T4c**<br>Privacy and data handling | **Definition.** HIPAA implications of AI tools that may retain or transmit protected health information, FERPA considerations in educational contexts, library and publisher contract restrictions on AI use with downloaded materials, secondary use of clinical data for AI training, and patient consent for AI processing of their data. Particularly salient for trainees who may feed patient information into consumer LLMs without recognizing the privacy implications.<br>Operational examples:<br>• Recognizing that pasting patient information into a public LLM without a Business Associate Agreement constitutes a HIPAA-relevant disclosure<br>• Understanding the privacy implications of an ambient AI scribe that processes patient encounter audio<br>• Following institutional policy regarding AI tools that may or may not be approved for clinical use |
| **T4d**<br>Ethics | **Definition.** Informed consent for AI involvement in care, equity considerations (subgroup performance disparities, access disparities), patient autonomy in the context of patient-facing AI, just deployment (responsible institutional adoption of AI tools, vendor selection, boundaries on when AI should NOT be used).<br>Operational examples:<br>• Recognizing when an AI tool may perform differently across patient demographic subgroups and adjusting reliance accordingly<br>• Considering whether AI-assisted care requires explicit patient consent or notification<br>• Participating in institutional decisions about AI tool selection or deprecation when patient safety or equity is at stake |

**Theme 5 · Cognitive Impacts of AI**

| Subtheme | Definition and operational examples |
|---|---|
| **T5a** Cognitive autonomy and deskilling [descriptor strengthened in v3] | **Definition.** Maintaining independent clinical reasoning when AI tools surface recommendations or differentials. Particularly relevant to trainees who may rely on AI before developing baseline clinical pattern recognition: risk of "neverskilling" (failing to develop competence in the first place, rather than losing established competence). For practicing physicians, the corresponding risk is deskilling (degradation of established competence with prolonged AI reliance).<br>Operational examples:<br>• Forming an independent clinical impression before consulting an AI tool for input<br>• Continuing to practice ECG interpretation skills even when AI ECG reads are routinely available<br>• Resisting the tendency to defer to AI-suggested workup when clinical judgment indicates a different course |
| **T5b** Anchoring and automation bias | **Definition.** Resisting the cognitive pull of AI-suggested differentials, disposition decisions, or risk scores when clinical judgment disagrees. Automation bias is the tendency to over-rely on AI recommendations even when they conflict with available evidence; anchoring bias is the tendency to fix on an initial AI-surfaced framing and fail to consider alternatives.<br>Operational examples:<br>• Avoiding premature closure on an AI-suggested diagnosis when the clinical picture does not fully fit<br>• Recognizing when an AI risk score is anchoring the team's disposition discussion in ways that don't reflect clinical judgment<br>• Maintaining diagnostic breadth when AI tools have narrowed focus prematurely |
| **T5c** Cognitive burden and alert fatigue | **Definition.** Managing the additional cognitive load of AI-generated alerts, popups, and recommendations alongside existing clinical decision support (CDS) systems. Includes recognition that AI alert fatigue can itself become a patient safety risk if it leads to dismissal of consequential alerts.<br>Operational examples:<br>• Triaging AI-generated alerts during a busy shift to attend to clinically significant ones<br>• Recognizing when AI alert volume is high enough to risk meaningful alerts being missed<br>• Contributing to institutional decisions about which AI alerts are worth the cognitive burden they impose |

**Table S2. Round 2 consensus statistics for all rated elements**

Importance ratings on a 5-point scale; n = 13. "% $\geq$ 4" is the proportion of panelists rating the element 4 or 5. A priori consensus required $\geq$ 70% rating $\geq$ 4 ($\geq$ 75% for themes) with interquartile range $\leq$ 1.

| Element | Mean | SD | % $\geq$ 4 | Status |
|---|---|---|---|---|
| **Derived competencies** | | | | |
| T1 derived competency | 4.36 | 0.50 | 100% | Consensus |
| T2 derived competency | 4.09 | 0.70 | 82% | Consensus |
| T3 derived competency | 4.45 | 0.52 | 100% | Consensus |
| T4 derived competency | 4.36 | 0.50 | 100% | Consensus |
| T5 derived competency | 4.27 | 0.65 | 91% | Consensus |

| Element | Mean | SD | % ≥ 4 | Status |
|---|---|---|---|---|
| **Themes** | | | | |
| T1 Communicating about AI | 4.18 | 0.75 | 91% | Consensus |
| T2 Understanding appropriate use cases | 3.91 | 0.83 | 91% | Consensus |
| T3 Interacting with AI | 4.55 | 0.52 | 100% | Consensus |
| T4 AI Risk Management | 4.27 | 0.65 | 91% | Consensus |
| T5 Cognitive Impacts of AI | 4.27 | 0.65 | 91% | Consensus |
| **Scenarios rated in Round 2** | | | | |
| S10 AI image interpretation | 4.09 | 0.70 | 82% | Consensus |
| S2+S15 AI-assisted documentation | 4.27 | 0.65 | 91% | Consensus |
| S11 Pre-emptive AI consultation | 4.18 | 0.60 | 91% | Consensus |
| S14 AI resource utilization | 4.27 | 0.65 | 91% | Consensus |
| S16 Patient-initiated AI | 4.36 | 0.50 | 91% | Consensus |
| S17 Post-hoc AI review | 3.82 | 0.98 | 55% | Borderline (retained, future-state) |
| Overall scenario-set adequacy | 4.18 | 0.75 | 91% | Consensus |

**Table S3. Architectural validation results (Round 2)**

| Architectural question | Result | Disposition |
|---|---|---|
| Is the one-to-one theme to competency mapping the right architecture? | 12 Yes · 1 No · 0 Revise | **Endorsed** |
| Rename Theme 2 to "Strengths and Limitations"? | 1 Yes · 9 No · 3 Maybe | **Rejected** |
| Is the Theme 4 risk-category structure preferable to a regulatory/governance structure? | 13 Yes · 0 No · 0 Maybe | **Unanimous** |
| Is "Cognitive Impacts of AI" the right name for Theme 5? | 12 Yes · 0 No · 1 Revise | **Endorsed** |

**Table S4. The ten clinical scenarios: full vignettes and consensus status**

Nine scenarios reached consensus; S17 was retained at borderline status with explicit future-state framing. Scenarios that passed clean Round 1 consensus (S1, S4, S6, S9) were not re-rated in Round 2.

| ID | Status | Vignette |
|---|---|---|
| **S1** | Passed (R1: 4.00, 75%) | **AI HPI summarization**<br>Vignette. AI generates an HPI summary from chart data (prior encounters, problem list, medication list); the physician reviews and edits before signing. Themes surfaced: T1a Documentation · T2b Limitations and weaknesses · T3c Evaluating AI evidence and output R1: mean 4.00, 75% ≥ 4, IQR 1.0 - passed · R2: Not re-rated; retained on confirmation (Section 2A) · Status: PASSED |
| **S4** | Passed (R1: 4.50, 100%) | **EKG reads Brugada**<br>Vignette. AI EKG interpretation flags a Brugada pattern; the physician evaluates the EKG, contextualizes against the clinical picture, and decides on workup and disposition. The AI-flagged finding may be visible to the patient via MyChart before clinician interpretation. Themes surfaced: T2b Limitations and weaknesses · T3a Workflow integration · T3c Evaluating AI evidence and output · T1c Patient-facing AI (via MyChart visibility) R1: mean 4.50, 100% ≥ 4, IQR 0.0 - passed (highest-rated R1 scenario) · R2: Not re-rated; retained on confirmation (Section 2A) · Status: PASSED |

| ID | Status | Vignette |
|---|---|---|
| **S6** | Passed (R1: 4.00, 75%) | **Stroke MCA localization** Vignette. AI imaging tool identifies a suspected MCA stroke and supports localization. The physician integrates AI output with the clinical exam to make decisions about acute treatment and disposition. Themes surfaced: T2b Limitations and weaknesses · T3a Workflow integration · T3c Evaluating AI evidence and output R1: mean 4.00, 75% $\geq$ 4, IQR 1.0 - passed · R2: Not re-rated; retained on confirmation (Section 2A) · Status: PASSED |
| **S9** | Passed (R1: 3.88, 75%) | **Smartwatch palpitations** Vignette. A patient arrives in the ED with smartwatch-flagged arrhythmia data (e.g., Smartwatch ECG showing atrial fibrillation). The physician integrates the wearable-derived AI interpretation into clinical reasoning and patient communication. Themes surfaced: T1c Patient-facing AI · T2a When AI is beneficial · T3c Evaluating AI evidence and output R1: mean 3.88, 75% $\geq$ 4, IQR 1.0 - passed · R2: Not re-rated; retained on confirmation (Section 2A) · Status: PASSED |
| **S10** | Passed (R2: 4.09, 82%) | **AI image interpretation** Vignette. The physician is reviewing imaging studies in the ED. An AI image-interpretation tool flags abnormalities (CXR findings, CT findings, ultrasound features) before or alongside physician review. The physician must adjudicate the AI's flags against personal interpretation and the clinical context. Themes surfaced: T2a When AI is beneficial · T2b Limitations and weaknesses · T3c Evaluating AI evidence and output · T5b Anchoring/automation bias R1: mean 3.62, 62% $\geq$ 4, IQR 1.0 - borderline · R2: mean 4.09, 82% $\geq$ 4, IQR 1.0 · Status: PASSED |
| **S2+15** | Passed (R2: 4.27, 91%) | **AI-assisted documentation** Vignette. AI tools generate clinical documentation either through ambient capture of the patient encounter (ambient AI scribes such as DAX Copilot) or through inline autocompletion of physician-typed notes. The physician must review, edit, and attest to the accuracy of the generated content before signing. Themes surfaced: T1a Documentation · T3a Workflow integration · T3c Evaluating AI evidence and output · T4a AI errors and failure modes R1: S15 ambient scribe mean 4.62, 88% (passed; top R1 scorer); S2 autocompletion mean 3.12, 50% (split) · R2: mean 4.27, 91% $\geq$ 4, IQR 1.0 · Status: PASSED Panel qualitative comments: ◦ "Two R2 panelists argued for splitting back: ambient scribes have layers and are largely only used in the patient care environment; autocompletion is distinct because it leads to a different cognitive process and deteriorating attention." ◦ "v3 keeps the combined scenario for parsimony (R2 quantitative endorsement remained strong) but the concept paper will acknowledge the distinction between the two cognitive risk profiles." |

| ID | Status | Vignette |
| --- | --- | --- |
| **S11** | Passed (R2: 4.18, 91%) | **Pre-emptive AI consultation**<br>Vignette. You are evaluating a patient in the ED. You consult an AI clinical decision support tool that uses the patient's presentation and medical record to suggest items to consider in the differential diagnosis. You must decide which of the AI's suggestions to incorporate into your reasoning and workup. Themes surfaced: T3a Workflow integration · T3c Evaluating AI evidence and output · T5a Cognitive autonomy and deskilling · T5b Anchoring/automation bias R1: Original popup framing: mean 4.00, 50% $\geq$ 4, IQR 2.0 (split) · R2: Active-consultation framing: mean 4.18, 91% $\geq$ 4, IQR 1.0 · Status: PASSED Panel qualitative comments: ∘ "S11 represents the operational reality of how trainees increasingly use tools like OpenEvidence. Paired with S17 to operationalize the pre-emptive vs post-hoc conceptual axis (§ 6.1)." |
| **S14** | Passed (R2: 4.27, 91%) | **AI resource utilization and flow**<br>Vignette. AI tools determine resource utilization and patient flow in your ED. Examples: AI selects which patient in the waiting room is brought back next; AI initiates lab ordering from triage data before physician evaluation; AI predicts disposition (admit vs discharge) at triage and adjusts downstream resourcing. You and your nursing colleagues must decide when to override AI's operational decisions. Themes surfaced: T3e Systems-level AI integration · T3f Interprofessional team-based AI · T2b Limitations and weaknesses R1: Original ESI triage scoring framing: mean 3.62, 62% $\geq$ 4, IQR 2.2 (broadest disagreement in R1) · R2: Resource utilization framing: mean 4.27, 91% $\geq$ 4, IQR 1.0 · Status: PASSED Panel qualitative comments: ∘ "One panelist noted: "And how to collect data on efficacy and counter-metrics for AI implementation; how to recognize drift, etc. Otherwise systems will tend towards the most frictionless experience even when models are ineffective." " |
| **S16** | Passed (R2: 4.36, 91%) | **Patient-initiated AI**<br>Vignette. An adult patient arrives in the ED with an acute presentation. Before evaluation, the patient has consulted a generative AI chatbot (such as ChatGPT, Claude, or Gemini) about their symptoms and arrives with an AI-generated differential or recommended workup. The physician must integrate the patient's AI-derived information into clinical reasoning and patient communication. Themes surfaced: T1b Risks and benefits with patients · T1c Patient-facing AI · T2a When AI is beneficial R2: mean 4.36, 91% $\geq$ 4, IQR 1.0 · Status: PASSED Post-R2 refinement: v3 incorporates three R2 panelist refinements: (1) narrowed scope to acute presentations (where the time pressure of patient-brought AI input is highest); (2) moved wearable-data scenarios to S9 (where they originally lived) to avoid overlap with the smartwatch palpitations scenario; (3) generalized "ChatGPT" to "chatbot" to avoid product specificity that will age poorly. Panel qualitative comments: ∘ "S16 is the scenario backing of T1c (patient-facing AI), together they address the most consistent qualitative signal from R1 free-text responses." |

| ID | Status | Vignette |
|---|---|---|
| **S17** | Borderline (R2: 3.82, 55%) - retained, future-state | **Post-hoc AI review**<br>Vignette. After completing your evaluation and initial documentation, an AI tool reviews your clinical note and reasoning. It surfaces potential omissions in your differential, missed considerations, or steps you may not have addressed. You decide whether to act on the AI's post-hoc suggestions. Themes surfaced: T3c Evaluating AI evidence and output · T5a Cognitive autonomy and deskilling (future-state framing) R2: mean 3.82, 55% $\geq$ 4, IQR 1.5 - borderline (did not meet consensus threshold) · Status: BORDERLINE Post-R2 refinement: v3 retains S17 with explicit future-state framing. R2 panel comment captured the disposition: "this is probably the future. May not be the highest priority." The pre-emptive vs post-hoc axis (§ 6.1) is conceptually important for the framework even if S17 is currently more aspirational than operational. The concept paper will frame S17 as the emerging integration model that prepares physicians for an alternative future state, paired with S11 which represents the current operational reality. Panel qualitative comments: ○ "Reference: Korom et al. 2025 (Penda Health AI Consult). The post-hoc model preserves clinical reasoning development; AI serves as a safety net rather than substitute for thinking." |

**Table S5. Decision audit trail (nominal group technique through version 3)**

| Phase | Decision | Resolution |
|---|---|---|
| **Phase 1 NGT** | Initial framework structure | 4 themes, 10 subthemes, and 14 scenarios consolidated from 19 candidate clinical scenarios; 7 candidate gap themes identified. |
| **Phase 1 to R1** | Pre-R1 addition | S15 (ambient AI scribe) added to the scenario set in early 2026 to reflect technology emergence between the NGT (spring 2025) and the Delphi launch. |
| **R1** | Scenario consensus | 5 scenarios reached consensus (S1, S4, S6, S9, S15); 9 did not, with varying splits. Strong qualitative signal that patient-facing AI was missing. All 4 themes reached adequacy. |
| **Meeting** | Decision 1 - scenario set | Dropped S3, S5, S7, S8, S12, S13. Kept S1, S4, S6, S9, S10. Combined S2+S15. Reframed S11 (post-hoc). Reworded S14 (resource flow). Added S16 (patient-initiated AI). Subtotal: 9 scenarios. |
| **Meeting** | Decision 2 - patient-facing AI | Both options adopted: a new T1c subtheme and a new scenario S16. |
| **Meeting** | Decision 3 - G6 disposition | Combined G6 (cognitive autonomy) and G1 (alert fatigue) into a new Theme 5, Cognitive Impacts of AI. |
| **Meeting** | Decision 4 - G4 + G5 disposition | Folded both into Theme 3 as new subthemes T3e (systems-level routing) and T3f (interprofessional). |
| **Meeting to v2** | Decision 5 - T4 substructure | Meeting set a 4-subtheme structure (Liability, Regulatory, Local governance, National governance) and renamed Theme 4 to "AI Risk Management." Co-leads then restructured to risk-category subthemes (Errors, Liability, Privacy, Ethics). R2 validation: risk-category structure unanimously endorsed (13/13). |
| **v2 (co-leads)** | Architecture | Articulated the one-to-one theme to competency mapping. The "critical appraisal" statement was repositioned as the Theme 2 derived competency rather than the framework's singular center. R2 validation: 12/13 endorsement. |

| Phase | Decision | Resolution |
|---|---|---|
| **v2 (co-leads)** | S11 restoration | S11 restored from the post-hoc reframe to a pre-emptive active-consultation framing reflecting current operational reality; S17 added as the paired post-hoc scenario. Both rated in R2. |
| **v2 (co-leads)** | T1c absorbs G2 | Former gap theme G2 (patient-generated data and wearables) folded into T1c, on the rationale that wearable interpretation requires AI and surfaces to patients first. |
| **R2** | Validation | All themes, all competencies, all subthemes, and 5 of 6 R2-rated scenarios reached consensus; S17 borderline. All architectural decisions endorsed. |
| **v3 (co-leads)** | Post-R2 refinements | Five refinements applied from R2 qualitative feedback: T2c reworded (Operating principles and decision logic); T3f scope extended (physician-APP/learner); T5a descriptor strengthened (deskilling/neverskilling); S16 narrowed (acute, no wearables, "chatbot" wording); S17 framed as future-state. |
